# Enhancing Radiation Oncology Training through Patient Advocate Integration

**DOI:** 10.1101/2025.07.28.25332319

**Authors:** Christina A. Simone-Soule, Sara E. Burke, Dema Abul-Enin, Charita Kunta, Adeseye Adekeye, Phuoc Tran, Amy Leader, Adam P. Dicker, Nicole L. Simone

## Abstract

**Purpose:** Patient advocates represent the voice of the patient community and bring a unique perspective to research. We hypothesized that including patient advocates in research training would increase trainees ability to communicate their science, understand the impact of their work and increase their empathy for patients.

**Methods:** An IRB-approved survey was administered to assess the impact of patient advocate feedback on trainees who presented their research via posters at our Radiation Oncology Research Symposium. Trainees reported demographics and self-assessed their use of lay language, changes in empathy, understanding the impact of their research, and potential future implications. The binomial proportion test and Fisher’s exact test were used to determine significance.

**Results:** The survey was completed by 80% (28/35) of trainees who participated in the poster session and interacted with patient advocates. Trainees were predominantly younger (60.7% under 30yo) and people of color (60.7%). Almost all trainees (96.4%) were comfortable talking to advocates but only 89.29% were comfortable using lay language. Trainees agreed (75%) that interacting with advocates increased their empathy. Most trainees (71.4%) believed patient advocates helped them understand the significance of their research and 64.3% believed advocates helped them develop new research ideas. Most trainees would like advocates at future poster presentations (85.7%), but did not want them to participate in study design or analysis. Gender and training level did not affect trainees’ ability to communicate in lay language, their empathy for patients, or understanding of their work’s clinical relevance.

**Conclusions:** Including patient advocates in poster sessions may improve trainees ability to present their research in lay language, increase their empathy and understand the clinical impact of their research. Future radiation oncology training should consider including the patient advocate voice to improve the tangible connection between research and real-world impact.

## Introduction

In oncology, integrating patient advocates into the research process has become increasingly recognized as a valuable practice^1^. These advocates bring essential patient and caregiver perspectives directly into the research process, ensuring that scientific advancements are patient-centered and clinically relevant. Their involvement is crucial in radiation oncology, where treatment decisions and technological innovations directly impact patient outcomes.

Patient advocates help humanize the research process and align it more closely with patient needs and experiences^2^ by participating in research design, implementation, and dissemination. This collaborative approach fosters a sense of urgency and purpose, as advocates represent the voices of those directly affected by cancer.

Despite the acknowledged importance of patient perspectives, many current training programs for radiation oncologists and researchers do not systematically include the patient voice^3^. This omission may lead to a gap in understanding the real-world implications of scientific research and clinical practice^4^. Incorporating patient advocates into educational settings, such as poster sessions and symposiums, can allow trainees to communicate their work in lay terms, enhance their empathy for patients, and better grasp the clinical significance of their research^5^. Such interactions improve communication skills and encourage a more holistic approach to scientific inquiry.

In this study, we hypothesized that the presence of patient advocates during radiation oncology poster sessions consisting of trainee presenters could enhance trainees empathy and understanding of clinical significance, and teach them to communicate with patients using lay language. We aimed to determine whether these interactions help trainees appreciate the broader impact of their work and facilitate better communication in non-technical language. This survey explores the potential benefits of including patient advocates in scientific presentations, ultimately contributing to more patient-centered research and clinical practices in radiation oncology.

## Methods and Materials

An IRB-approved survey was administered to trainees who presented their research via posters in a poster symposium in a Radiation Oncology Department at a NCI Designated Comprehensive Cancer Center and interacted with patient advocates to present their research. Trainees were defined by the Cancer Research Training and Education Coordination definition to include individuals at various stages of their education and careers, including high school and college students, graduate students, medical students, postdoctoral fellows, clinical residents and fellows, and junior faculty. The trainees created posters of their radiation-related research and presented them at a poster symposium that was attended by invited patient advocates, department employees and faculty at all levels, research teams and cancer center leadership.

Patient advocates were former patients from the department who expressed to the treating physician they would like to volunteer in the department. They received guidance before the event on how to engage with trainees. They were instructed to visit each poster, ask open-ended questions to understand the research, seek clarification when needed, and share personal insights about how the research might impact future patients. The Department of Radiation Oncology poster symposiums of 2023 and 2024 incorporated the patient advocates who had a checklist to attend every trainee poster. To assess this interaction, a novel survey was developed to assess the perceived impact of patient advocates on presenters’ communication skills, understanding of clinical significance, and engagement with patient advocacy. The survey included 10 questions across several domains: (1) demographic information, (2) comfort in discussing research with patient advocates, (3) ability to communicate research in lay language, (4) perceived empathy and clinical relevance gained from advocate interactions, (5) influence of advocates on research interpretation and future involvement in advocacy, and (6) preferences for advocate participation in future symposia and research activities (Supplemental Table 1). A 5-point Likert scale ranging from “Strongly Agree” to “Strongly Disagree,” and an open-ended section allowed for additional feedback on improving the integration of patient advocates in research discussions. The anonymous survey included a consent statement, and did not collect protected health information. No compensation was provided for participation.

Survey responses from Likert scale questions were combined into two groups for comparison: “strongly disagree,” “disagree,” and “neutral,” and “agree,” and “strongly agree”. A binomial test for proportions was used to assess statistical significance and the hypothesized proportion was set to 0.5. Fisher’s exact test was applied to the survey questions when comparing the survey answers between two different demographics groups. Statistics were completed using R Studio Version 4.3.1.

## Results

Of the 35 trainees in 2023 and 2024, 80% (n=28) completed the survey. Of the respondents, 50% identified as female, and 60.7% were younger than 30, 60.7% identified as People of Color. The trainees included 3.6% high school students, 35.7% graduate or medical students, 21.43% medical or physics residents, or postdoctoral fellows, 14.3% junior faculty, and 14.3% were post-baccalaureate students/lab assistants (Table 1).

**Table 1.** Presenter Characteristics.

| **Table 1.** | **% Responses (n)** |
| --- | --- |
| **Age (yo)** |  |
| <30 | 64.29 (18) |
| ≥31 | 35.71 (10) |
| **Gender** |  |
| Male | 50 (14) |
| Female | 50 (14) |
| **Race/Ethnicity** |  |
| Asian | 21.43 (6) |
| Black | 25 (7) |
| Multiracial/Multiethnic | 7.14 (2) |
| Hispanic or Latino | 7.14 (2) |
| White | 39.29 (11) |
| **Poster Presenter's Position** |  |
| High School Student | 3.57 (1) |
| Graduate/Medical Student | 35.71 (10) |
| Medical/Physics Residents | 21.43 (6) |
| Postdoctoral Fellows | 10.71 (3) |
| Postbaccalaureate Lab Assistants | 14.29 (4) |
| Junior Faculty | 14.29 (4) |

### Effect of Patient Advocates on Communication, Empathy, and Research Relevance

The majority of trainees reported beneficial interactions with advocates. Overall, 96.4% of the trainees agreed or strongly agreed that they were comfortable talking to advocates (p<0.01). 89.29% of participants strongly agreed or agreed that they were able to talk to the patient advocate in lay language (p<0.01). The inclusion of patient advocates increased presenters’ empathy, with 75% agreeing or strongly agreeing that these interactions enhanced their understanding of patients’ experiences (p=0.01). The majority of poster presenters (71.4%) believed that patient advocates helped them understand the clinical significance of their research (p<0.05), while 64.3% felt advocates contributed ideas to complement their work (p=0.09). However, half of the respondents did not think advocates could help them identify novel conclusions (p=0.575). (**Figure 1**).

**Figure 1.**
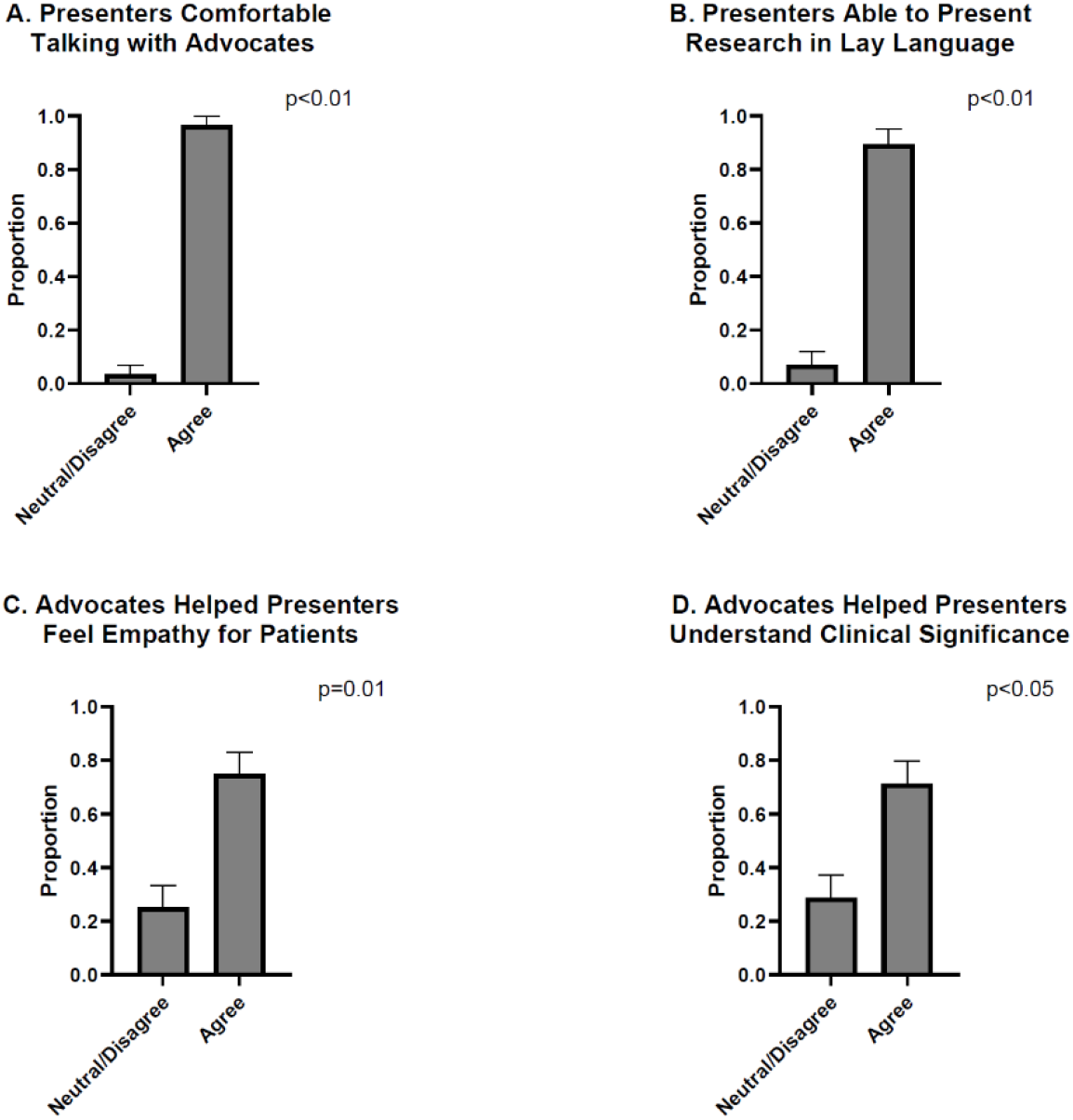
Effect of Patient Advocates on Communication, Empathy, and Research Relevance. A. 96.4% of respondents agreed or strongly agreed that they felt comfortable communicating with advocates (p<0.01). B. Additionally, 89.29% of participants agreed or strongly agreed that they could converse with the patient advocate using lay language (p<0.01). C. Respondents indicated that patient advocates enhanced presenters’ empathy, with 75% agreeing or strongly agreeing that these interactions deepened their understanding of patients’ experiences (p=0.01). D. Most poster presenters (71.4%) believed that patient advocates helped them grasp the clinical relevance of their research (p<0.05). E. Additionally, 64.3% felt that advocates contributed ideas that complemented their work (p=0.095). F. Roughly half of the respondents did not believe advocates played a role in identifying novel conclusions (p=0.575).

### Trainee Interest in Future Advocate Engagement

The survey also evaluated the interest of trainees in future patient advocacy initiatives. A majority of trainees (78.6%) strongly agreed or agreed that they would like to engage in future patient advocacy efforts (p<0.01), highlighting a strong interest in maintaining patient-centered elements in research training. Additionally, 85.7% of trainees expressed a desire for patient advocates to be present at future poster sessions (p<0.01), reinforcing the perceived value of their participation. However, respondents were less confident about the role of advocates in study design or data analysis, with 67.86% of trainees stating that they would like the patient advocates to participate in study design (p=0.043) and only 46.43% stating that they would like the patient advocates to participate in data analysis (p=0.714), suggesting that while patient input is valued in communication and application, there is hesitation about integrating patient voices into technical aspects of research (**Figure 2**).

**Figure 2.**
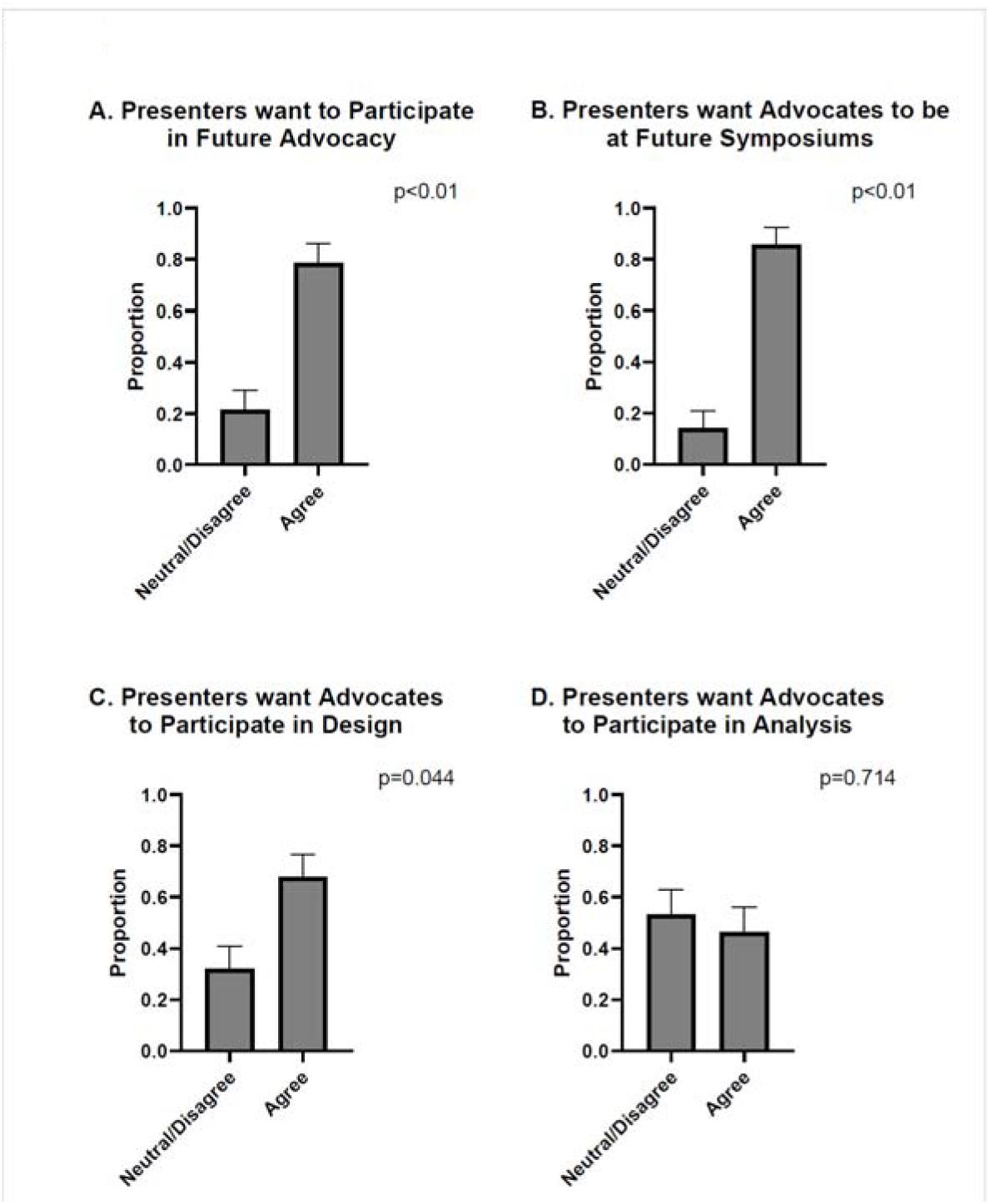
Trainee Interest in Future Engagement with Patient Advocates. A. The majority of respondents (78.6%) agreed or strongly agreed that they would like to take part in future patient advocacy efforts (p<0.01), emphasizing a strong commitment to integrating patient-centered elements into research training. B. Furthermore, 85.7% of participants expressed interest in having patient advocates present at future poster sessions (p<0.01), underscoring the perceived benefits of their involvement. C. However, respondents were more uncertain about the advocates’ role in study design and data analysis. While 67.86% indicated an interest in having patient advocates contribute to study design (p=0.044), D. only 46.43% supported their involvement in data analysis (p=0.714).

### Differences in Responses by Demographics

When examining responses within different demographic groups, no differences emerged in perceptions of advocate interactions. Both women and men, and students and other trainees, agreed or strongly agreed that they could effectively communicate their research in lay terms (p=ns). Similarly, no differences were observed in self-reported empathy with both genders and all training levels having reporting the same increase in empathy for patients following the advocate interaction (p=ns). (Figure 3).

**Figure 3.**
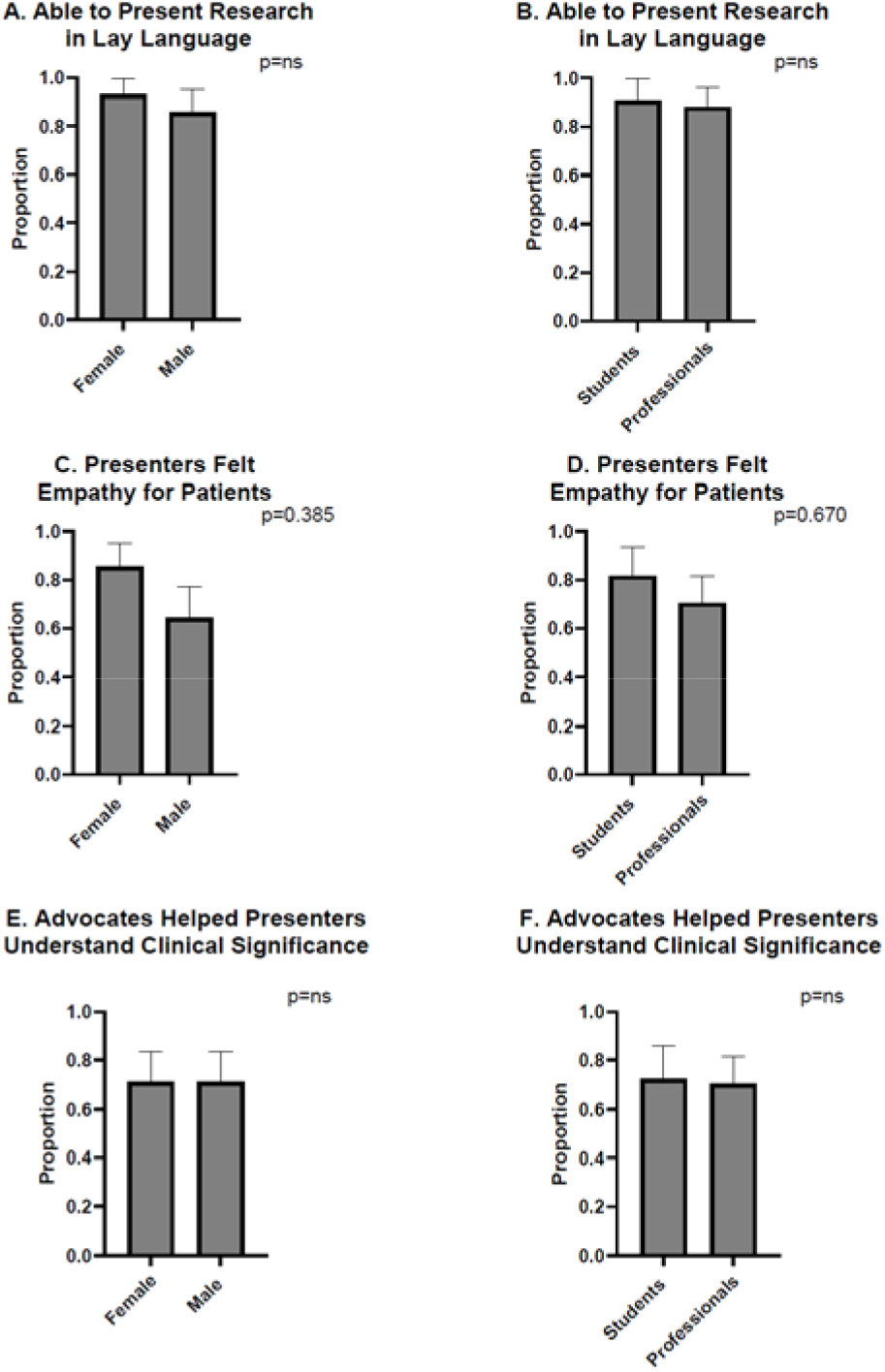
Response to patient advocate interaction was similar between trainees of different genders and training levels. Women and men equally agreed they could effectively explain their research in lay terms (p=ns) just as did trainees who were students or postgraduates/junior faculty (p=ns). B. No differences were noted in self-reported empathy between genders (C) (p=0.385) or level of training (D) (p=0.670). Similarly neither gender (E) or training level (F) influenced the trainees ability to grasp the clinical relevance of their work compared to non-students (p=ns).

## Discussion

This study highlights the significant impact of patient advocates on the educational experience of trainees in radiation oncology. The findings indicate that patient advocates during poster sessions enhanced trainees’ ability to communicate their research in lay language, deepened their empathy for patients, and improved their understanding of the clinical significance of their work. Furthermore, trainees expressed a strong interest in incorporating patient advocates into future research endeavors, emphasizing the potential for long-term collaboration. The results also revealed differences in engagement levels across subpopulations, providing insights into how diverse groups interact with advocacy initiatives.

A key finding of this study was that trainees reported increased confidence in translating complex scientific concepts into language accessible to non-expert audiences. Communicating in lay terms is critical for effective patient-provider interactions and broader public engagement in science^6^. Our results align with prior studies demonstrating that patient advocate involvement enhances science communication skills among researchers^7^. In particular, research in oncology education has shown that exposure to patient perspectives fosters more patient-centered communication strategies, reinforcing the importance of including advocacy in training programs^8^.

In addition to enhancing communication skills, trainees recognized the value of continued collaboration with patient advocates. Many trainees indicated that they would welcome advocate involvement in future poster sessions, research design and data interpretation. These findings mirror previous studies in which patient advocates contributed meaningful insights to clinical and translational research projects, thereby improving study relevance and applicability^9^. Since the benefit of discussing research with patient advocates did not differ between training level or gender of trainee, educational initiatives may be beneficial to ensure that all trainees can effectively engage with and learn from patient advocates.

This study had limitations. The relatively small sample size may limit the generalizability of the findings, and self-reported data introduces potential biases. The study did not assess long-term retention, leaving the durability of communication and empathy gains unknown. Future research should explore lasting impacts, formal curriculum integration, and use larger samples with objective measures,

This study underscores the transformative value of integrating patient advocates into radiation oncology training. By fostering empathy, sharpening communication, and reinforcing the clinical relevance of research, their presence can help shape a more patient-centered generation of scientists and clinicians in the field of radiation oncology.

## Conclusions

This study provides compelling early evidence that integrating trained patient advocates into radiation oncology training is both feasible and transformative. While patient-centered research has gained traction in other areas of oncology, our findings demonstrate for the first time that involving patient advocates in trainee research presentations significantly enhances communication skills, empathy, and clinical relevance—three competencies foundational to effective oncologic care and innovation. Importantly, the enthusiasm expressed by trainees to continue advocate engagement suggests that this model not only improves immediate educational outcomes, but may also instill enduring values of partnership and patient-centeredness. These results position patient advocates not as passive bystanders but as active contributors to research culture and education. As radiation oncology grapples with growing technological complexity and patient distrust, embedding the patient voice into training is a timely and scalable intervention. Future studies should explore longitudinal outcomes, integrate this model across multiple institutions, and assess whether advocate engagement during training ultimately translates to more humanistic, impactful clinical and research careers.

## Data Availability

All data produced in the present study are available upon reasonable request to the authors.

## Data Availability Statement

Research data are stored in an institutional repository and will be shared upon request to the corresponding author

## Acknowledgements

We would like to thank the amazing patient advocates who have dedicated their perspective and time to help train our Radiation Oncology trainees.

